# Clinical characteristics and outcome of immunocompromised patients with COVID-19 caused by the Omicron variant: a prospective observational study

**DOI:** 10.1101/2022.04.25.22273197

**Authors:** S. Reshwan K. Malahe, Rogier A.S. Hoek, Virgil A.S.H. Dalm, Annoek E.C. Broers, Caroline M. den Hoed, Olivier C. Manintveld, Carla C. Baan, Charlotte M. van Deuzen, Grigorios Papageorgiou, Hannelore I. Bax, Jeroen J. Van Kampen, Merel E. Hellemons, Marcia M.L. Kho, Rory D. de Vries, Richard Molenkamp, Marlies E.J. Reinders, Bart J.A. Rijnders

## Abstract

**Background:** In the general population, illness after infection with the SARS-CoV-2 Omicron variant is less severe compared with previous variants. Data on the disease burden of Omicron in immunocompromised patients are lacking. We investigated the clinical characteristics and outcome of a cohort of immunocompromised patients with COVID-19 caused by Omicron.

**Methods:** Solid organ transplant recipients, patients on anti-CD20 therapy, and allogenic hematopoietic stem cell transplantation recipients on immunosuppressive therapy infected with the Omicron variant, were included. Patients were contacted regularly until symptom resolution. Clinical characteristics of consenting patients were collected through their electronic patient files. To identify possible risk factors for hospitalization, a univariate logistic analysis was performed.

**Results:** A total of 114 consecutive immunocompromised patients were enrolled. Eighty-nine percent had previously received three mRNA vaccinations. While only one patient died, 23 (20%) required hospital admission for a median of 11 days. A low SARS-CoV-2 IgG antibody response (<300 BAU/mL) at diagnosis, higher age, being a lung transplant recipient, more comorbidities and a higher frailty were associated with hospital admission (all p<0.01). At the end of follow-up, 25% had still not fully recovered. Of the 23 hospitalized patients, 70% had a negative and 92% a low IgG (<300 BAU/mL) antibody response at admission. Sotrovimab was administered to 17 of them, of which one died.

**Conclusions:** While the mortality in immunocompromised patients infected with Omicron was low, hospital admission was frequent and the duration of symptoms often prolonged. Besides vaccination, other interventions are needed to limit the morbidity from COVID-19 in immunocompromised patients.

**Summary:** COVID-19-associated morbidity and mortality in immunocompromised patients is unknown for the SARS-CoV-2 Omicron variant. This prospective registry, demonstrated low COVID-19-associated mortality in these vulnerable patients. However, morbidity remained substantial. Other interventions to abate COVID-19 severity are needed.

## INTRODUCTION

Solid organ transplant recipients (SOTR), patients treated with B-cell depleting therapy, and allogenic hematopoietic stem cell transplantation recipients, are at increased risk of severe COVID-19-associated morbidity and mortality [1–4]. Several comorbidities that were previously associated with more severe COVID-19 are frequently present in immunocompromised patients and the use of immunosuppressive agents further increases the risk of a poor outcome. Although vaccination effectively protects against severe COVID-19 disease in the general population, the humoral and cellular immune response after vaccination of immunocompromised patients is lower, and protection from disease therefore reduced [5,6].

In November 2021, the SARS-CoV-2 Omicron variant emerged and was rapidly declared as variant of concern (VOC) by the World Health Organization (WHO). The high number of mutations observed in the spike (S) protein reduces or completely abolishes the neutralizing capacity of antibodies induced by previous infection, or after a standard vaccination regimen [7]. Furthermore, the Omicron variant is more infectious than any previous VOC.^7^ In addition to evading pre-existing immunity, most monoclonal antibody therapies are ineffective against the Omicron variant [8]. Sotrovimab is the only approved monoclonal antibody that retained activity against the Omicron BA.1 variant [9]. Currently a second sublineage of the Omicron variant (BA.2) is rapidly replacing BA.1 in most parts of the world and a reduced in vitro activity of sotrovimab against Omicron BA.2 is observed [10,11].

Infections with the Omicron variant have been associated with diminished morbidity and mortality [12]. To some extent, this can be explained by other immunological correlates, like cross-reactivity of vaccination-or infection-induced virus-specific T-cells [13,14]. Furthermore, animal models suggest a change in the pathophysiology of Omicron, with a shift towards infection of the upper rather than the lower airways [15]. The uncoupling of the extremely high incidence of Omicron infections and only a moderate increase in hospital and ICU admissions, has led several countries to loosen or completely stop public health measures previously implemented. It is therefore unpreventable that Omicron will circulate across the population, leading to exposure of immunocompromised patients to this variant. To date, no data are available on the clinical course and outcome of COVID-19 caused by Omicron in immunocompromised patients, in which protection from preceding infections or vaccinations is probably reduced. Here, we report on the morbidity and mortality of Omicron infections in immunocompromised patients in care at the Erasmus University Medical Center in the Netherlands.

## METHODS

Approximately 200 kidney, 70 liver, 35 lung, and 15 heart transplants as well as 100 allogeneic hematopoietic stem cell transplantations (alloHSCT) are performed annually at Erasmus MC. A prospective registry was implemented of SOT recipients and patients from the department of hematology and internal medicine (clinical immunology), infected with the Omicron variant. Patients had been instructed to contact their treating physician when a diagnosis of COVID-19 was suspected or confirmed (e.g. self-test or at a public health testing location). Kidney and lung transplant recipients with a positive self-test were invited at the hospital to collect a home monitoring kit, that included an oxygen saturation meter and instructions on whom to contact when saturation level declined to 93% or lower. All patients that reported a positive test were instructed to re-contact their treating physician if symptoms did not abate within five to seven days, or when symptoms worsened. In addition, all patients were re-contacted proactively up until March 14, 2022. Patients that were included fulfilled the following inclusion criteria: i) a proven SARS-CoV-2 infection with the Omicron variant in the period between December 13, 2021 and February 3, 2022 or a SARS-CoV-2 infection confirmed by polymerase chain reaction (PCR)-based assay or an antigen self-test after the 9th of January 2022 (when Omicron had become >95% dominant in the Netherlands), ii) SOTR (kidney, liver, lung, heart, or multi-organ) or immunocompromised due to the use of anti-CD20 therapy for an auto-immune or hematological disease, or an alloHSCT recipient on immunosuppressive therapy for the prevention or treatment of graft versus host disease (GVHD), and iii) follow-up of at least two weeks after the onset of symptoms.

### Data Collection

Demographic data, medical history, comorbidities, medication use, vaccination status, route to diagnosis (in-or outpatient), hospitalization status, clinical features, antibody titer before and after diagnosis (LIAISON ® SARS-CoV-2 Trimeric Spike IgG), treatment data, and the clinical outcome, were collected from all patients from the electronic patient files. Clinical Frailty Scale (CFS) was used to score the health status of patients by using the clinical assessment of the Canadian study of Health and Ageing [16]. To evaluate symptom duration, all SARS-CoV-2 infected patients were contacted by telephone up to March 14, 2022.

The SARS-CoV-2 variant and sublineage were determined by detection of variant-specific single-nucleotide polymorphisms (SNPs), including K417, S371 and S373, using PCR and melting-curve analysis (VirSNiP 53-0787-96 and 53-0827-96, TIBMOLBIOL, Berlin, Germany). More information about this SNP analysis is available in the online supplement.

For patients who did not perform PCR testing at the Erasmus MC, SARS-CoV-2 infection was diagnosed at the Municipal Health Service (MHS) by reverse transcriptase (RT)-PCR or by means of an antigen self-test, and the variant could therefore not be determined. These patients were only included when the onset of symptoms started after January 9, 2022, the date from which the Omicron VOC had become responsible for >95% of the SARS-CoV-2 infections in the Netherlands [17].

### Standard Immunosuppressive Regime

Information about the immunosuppressive therapies and regimens used in the different organ transplant recipients is available in the online supplement.

### COVID-19 Treatment

Hospitalized COVID-19 patients were treated according to the Dutch COVID-19 guideline, which includes dexamethasone for all patients requiring supplemental oxygen, tocilizumab for those with a CRP >74 mg/L and at least 6 liter oxygen/min [18]. Sotrovimab became available on January 24, 2022 in the Netherlands. As the natural course of Omicron in immunocompromised patients was unclear at that time, the vaccination coverage high, and the capacity to treat outpatients with sotrovimab limited, it was not implemented as outpatient therapy. However, sotrovimab was used for all immunocompromised patients that required hospital admission as soon as it became available (500 mg intravenously upon admission).

### Dutch COVID-19 Vaccination Strategy

According to Dutch guidelines, all SOTR, B-cell depleted and alloHSCT patients, are considered fully vaccinated against COVID-19 when they received three doses of an mRNA-based vaccine. This is based on the low response of neutralizing antibodies after two vaccinations in these groups. The additional fourth dose is then considered as booster [19].

### Statistical Analysis

Analyses were performed in SPSS (version 28) and GraphPad Prism 9. Continuous data are presented as mean with standard deviation (SD) or as median with interquartile range (IQR) in case of non-normal distribution. Categorical data are presented as percentages. The primary outcome was hospitalization during the follow-up period. Since this study was neither powered nor designed to identify independent risk factors for hospitalization, analyses were purely explorative. To identify possible independent risk factors for hospitalization, the associations of patient characteristics with risk of hospitalization were examined using univariate logistic regression analysis. Odds ratios (ORs), corresponding 95% confidence intervals (95% CIs) and p-values were reported. A p-value of <0.05 was considered statistically significant. A cut-off level of ≤300 BAU/mL for appropriate vaccination response, is based on a study that explored the association between SARS-CoV-2-binding antibody concentration and neutralizing antibody titer against the D614G variant [20].

### Ethical Approvals

The institutional review board at Erasmus MC University Medical Center in Rotterdam (officially called “Medisch Ethische Toetsingscommissie Erasmus MC, METC Erasmus MC”) reviewed the protocol. They confirmed that this study does not fall under the Dutch law on research in human subjects (WMO) and therefore waived the requirement for ethical approval of this study. However, all SOTR provided written informed consent for the use of their clinical data as part of an ongoing quality improvement program and the non-SOTR group consented for use of their data in the context of this study.

## RESULTS

### Demographics and Baseline Characteristics

A total of 114 immunocompromised patients with a SARS-CoV-2 infection caused by Omicron, were included in the period between December 2021 and February 2022, and were followed for at least two weeks after onset of symptoms. Of these, 100 were SOTRs and 14 were immunocompromised as a result of anti-CD20 therapy for auto-immune or hematological diseases, or because they were receiving immunosuppressive therapy after alloHSCT. Of the SOTRs, 43 were kidney, 16 lung, 19 liver, 17 heart, and five multi-organ transplant recipients. The demographics and baseline characteristics of the patient cohort are listed in Table 1.

**Table 1:**
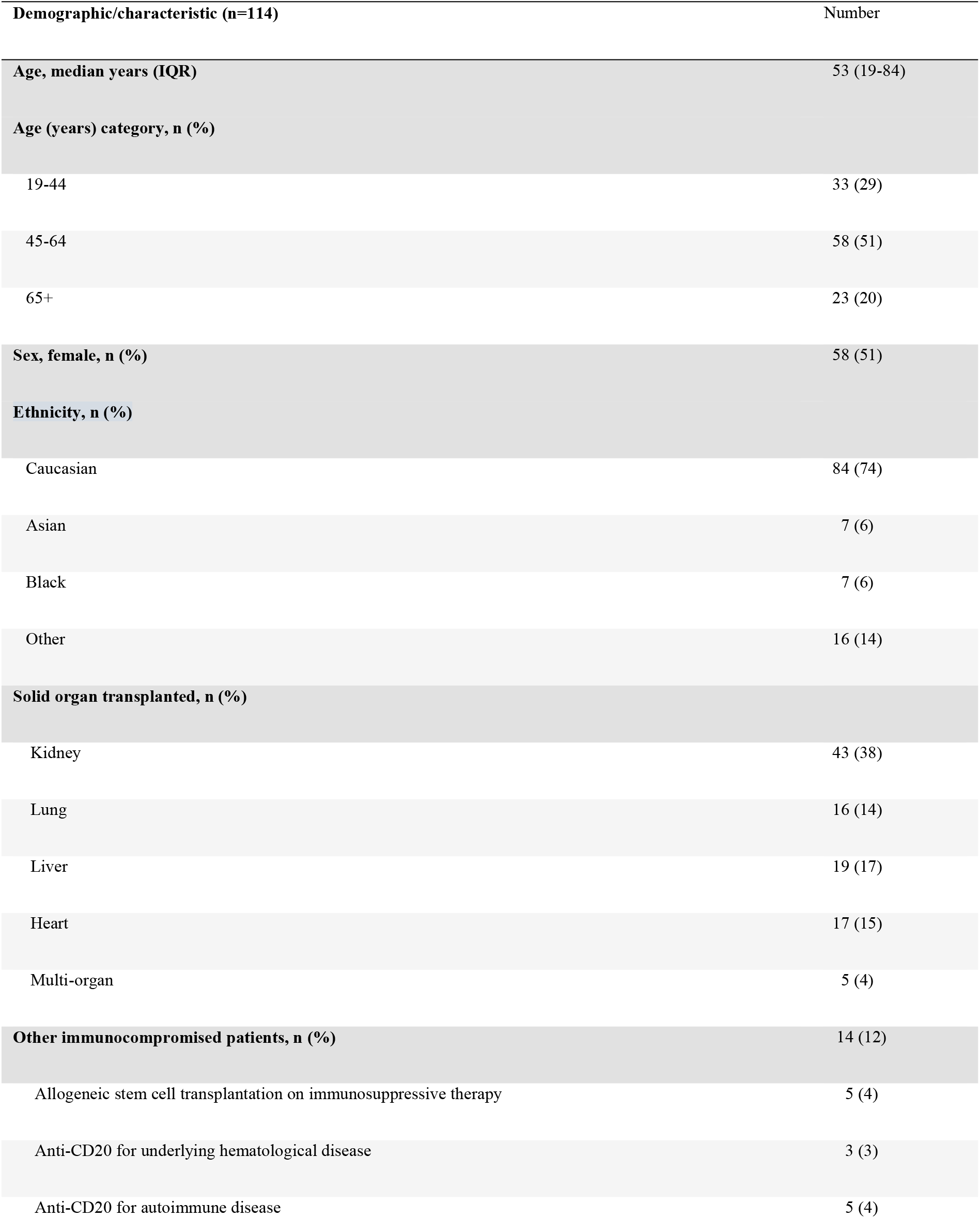
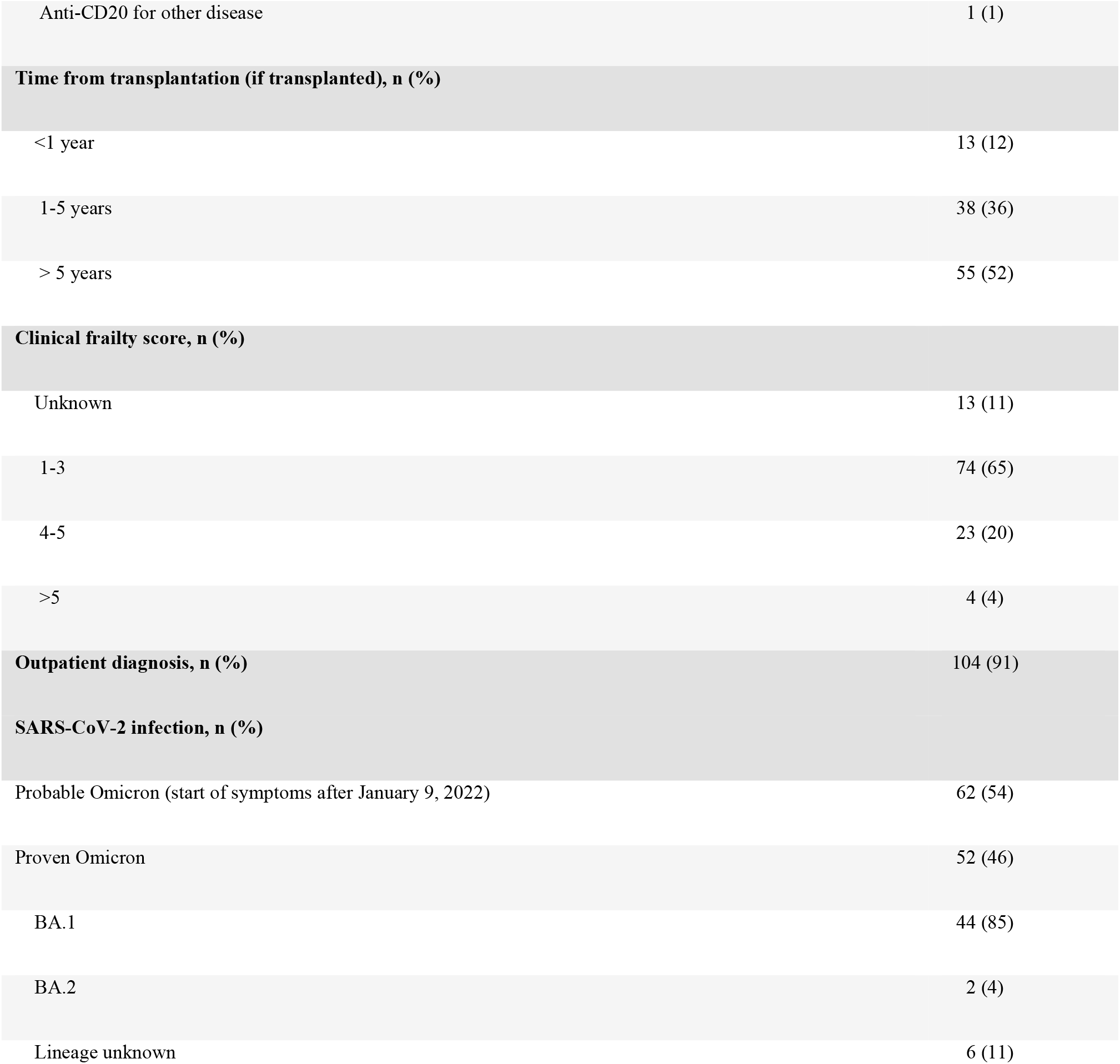
Patient demographics.

Median age of the immunocompromised patients was 53 years. Half of the patients were female and most of them were Caucasian. Of the transplant recipients, 52% received their transplant more than five years ago. Most of the patients’ health was scored as very fit, well or managing well, and most of them were diagnosed with COVID-19 in the outpatient setting. Omicron was proven to be the causative variant in 46% of the SARS-CoV-2 infections, in which 85% was caused by the BA.1 variant. In the remaining 54% of the SARS-CoV-2 infections, the diagnosis was made at the MHS by RT-PCR or by means of an antigen self-test. In these cases, symptom onset started in the period when the Omicron variant was responsible for at least 95% of SARS-CoV-2 infections in the Netherlands, and therefore considered Omicron [17]. The most reported symptoms included: rhinitis (54%), cough (53%), malaise (46%), fever (40%) headache (40%), sore throat (36%), fatigue (22%), gastro-intestinal complaints (14%), and myalgia (10%). Only one patient was asymptomatic during the complete follow-up period. The most frequent comorbidities were: arterial hypertension (65%), chronic kidney disease defined as eGFR <60 mL/min/1.73 m^2^ (29%), diabetes mellitus type 2 (28%), and atherosclerotic cardiovascular disease or heart failure (15%).

### Clinical Features and Outcome

The clinical features and outcome in the non-hospitalized and hospitalized immunocompromised patients are listed in Table 2. Among 23 hospitalized immunocompromised patients with a median age of 63 years, 57% were male. Among these patients, 35% required supplemental oxygen and 30% required high-flow nasal cannula therapy. Seventy-four percent received sotrovimab, 74% dexamethasone, 35% anti-IL6 therapy, and 13% methylprednisolone pulse therapy. None of the patients required mechanical ventilation. The median duration of hospitalization was 11 days (range 2-25 days) and 11 patients (48%) were hospitalized for more than ten days. In 65% of the hospitalized patients, the COVID-19 diagnosis had been made at least 48 hours before hospital admission. The majority (70%) was seronegative at the time of hospital admission, despite the fact that 78% had been fully vaccinated. Only five hospitalized patients (22%) had received their booster (i.e. fourth) vaccination. One hospitalized patient died. This was a lung transplant recipient with multiple comorbidities, in whom a “do not ventilate” decision had been made. Figure 1 illustrates the time to full-symptom resolution. The median duration of symptoms was 14 days and at the end of the follow-up period (68 days after onset of symptoms), 25% had still not fully recovered.

**Table 2:** Clinical features and outcomes in non-hospitalized and hospitalized patients.

|  | Non-hospitalized | Hospitalized |
| --- | --- | --- |
| **Number, n (%)** | 91 (80) | 23 (20) |
| **Age, median years (IQR)** | 50 (19-84) | 63 (42-74) |
| **Age (years) category, n (%)** |  |  |
| 19-44 | 32 (35) | 1 (4) |
| 45-64 | 48 (53) | 11 (48) |
| 65+ | 11 (12) | 11 (48) |
| **Sex, male, n (%)** | 43 (47) | 13 (57) |
| **Ethnicity, non-Caucasian, n (%)** | 21 (23) | 9 (39) |
| **Solid organ transplant recipient, n (%)** |  |  |
| Kidney | 34 (37) | 9 (39) |
| Lung | 5 (6) | 11 (48) |
| Liver | 19 (21) | 0 |
| Heart | 17 (19) | 0 |
| Multi-organ | 4 (4) | 1 (4) |
| **Other immunocompromised patients, n (%)** | 12 (13) | 2 (9) |
| **Time from transplantation (if transplanted)** |  |  |
| < 1 year | 10 (11) | 3 (13) |
| 1-5 years | 29 (32) | 9 (39) |
| >5 years | 46 (51) | 9 (39) |
| **Maintenance immunosuppression, n (%)** |  |  |
| CNI monotherapy | 14 (15) | NA |
| CNI + MMF | 27 (30) | 5 (22) |
| Pred + CNI | 9 (10) | NA |
| Pred + CNI + MMF | 14 (15) | 11 (48) |
| Pred + CNI + MMF + EVE | NA | 2 (9) |
| Anti-CD20 monotherapy | 8 (9) | 0 |
| Other<sup>a</sup> | 19 (21) | 5 (22) |
| Immunosuppression containing MMF | 51 (56) | 18 (78) |
| **Positive PCR ≥ 48 hours before hospitalization, n (%)** | NA | 15 (65) |
| **COVID-19 treatment, n (%)** | NA |  |
| No oxygen requirement |  | 8 (35) |
| 1-5 liters/min oxygen |  | 6 (26) |
| 5-15 liters/min oxygen |  | 2 (9) |
| High-flow nasal cannula therapy |  | 7 (30) |
| Mechanical ventilation |  | 0 |
| Sotrovimab treatment |  | 17 (74) |
| Dexamethasone treatment |  | 17 (74) |
| Anti-IL6 treatment |  | 8 (35) |
| Methylprednisolone pulse<sup>b</sup> |  | 3 (13) |
| **Time to hospitalization, median days (IQR)** | NA | 10 (1-34) |
| **Duration of hospitalization, median days (IQR)** | NA | 11 (2-25) |
| **Duration of hospitalization, n (%)** | NA |  |
| 2-5 days |  | 6 (26) |
| 6-10 days |  | 6 (26) |
| >10 days |  | 11 (48) |
| **Clinical outcome at the end of follow-up, n (%)** |  |  |
| Not recovered | 16 (18) | 12 (52) |
| Recovered | 75 (82) | 10 (44) |
| Death | 0 | 1 (4) |
| **Time to recovery (if recovered), median days (IQR)** | 10 (2-49) | 22 (4-44) |
| **SARS-CoV-2 spike IgG (BAU/mL), mean titer (SD)<sup>c</sup>** | 3771 (8466) | 46 (96) |
| **SARS-CoV-2 spike IgG category (BAU/mL), n (%)** |  |  |
| Seronegative | 19 (21) | 16 (70) |
| Seropositive (ELISA)<sup>d</sup> | 6 (7) | NA |
| 33.8-299<sup>e</sup> | 1 (1) | 5 (22) |
| 300-1000 | 5 (5) | 1 (4) |
| >1000 | 12 (13) | 0 |
| Unknown | 48 (53) | 1 (4) |
| **Vaccination status, n (%)** |  |  |
| Non vaccinated | 8 (9) | 5 (22) |
| Fully vaccinated | 83 (91) | 18 (78) |
| Boosted | 9 (10) | 5 (22) |

Abbreviations: CNI= calcineurin inhibitor, MMF= mycophenolate mofetil, Pred= prednisolone, PCR= polymerase chain reaction, IgG= immunoglobulin G, ELISA= enzyme-linked immunosorbent assay, EVE= everolimus, ADM= adalimumab, SRL= sirolimus, CsA= ciclosporin A, AZA= azathioprine, TCZ= tocilizumab, Bela= belatacept, ETN= etanercept
<sup>a</sup>: Other immunosuppressive regimens include: Pred + CNI + EVE (n=3), Pred + CNI + ADM (n=1), CNI + ADM (n=1), MMF + SRL (n=1), CNI + EVE (n=1), MMF mono (n=1), Pred + SRL (n=1), Pred + MMF (n=1), CsA mono (n=1), an oncolytic (n=1), Pred mono (n=1), Pred + CNI + AZA (n=1), Pred + AZA (n=1), CNI + AZA (n=1), Pred + MMF + CsA (n=2), Pred + CsA (n=2), Pred + CNI + MMF + TCZ (n=1), Pred + MMF + Bela (n=2), CNI + MMF + ETN (n=1)
<sup>b</sup>: On top of or after dexamethasone
<sup>c</sup>: If SARS-CoV-2 spike IgG is known
<sup>d</sup>: In some patients, only an ELISA was performed to detect SARS-CoV-2 seroconversion, which only gives a positive or negative result and no IgG titer
<sup>e</sup>: 33.8 BAU/mL is the detection limit of the LIAISON test

**Figure 1:**
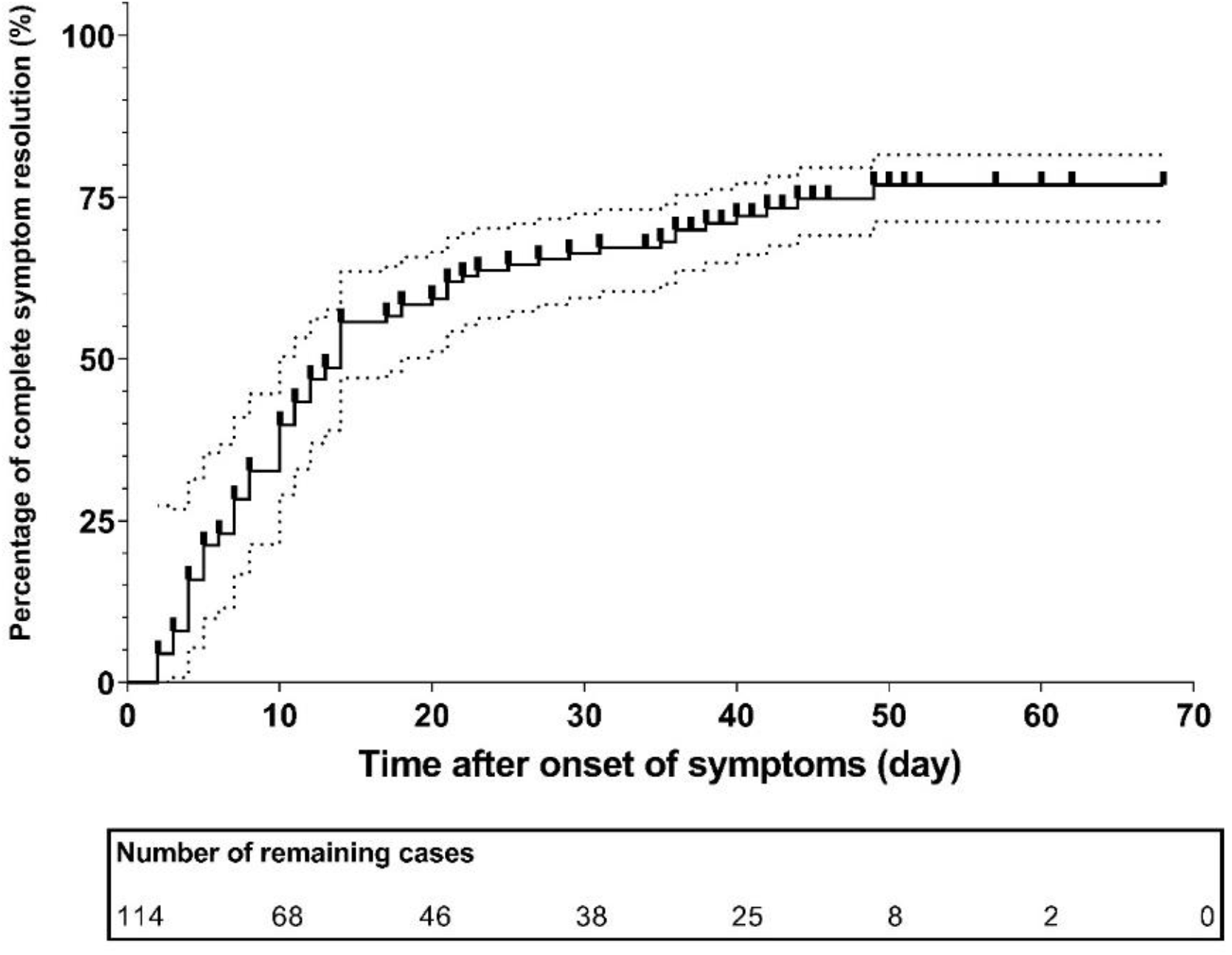
Time to symptom resolution in all 114 immunocompromised patients.

Based on evolving observation that the hospital admission rate of the lung transplant recipients was very high compared to all other immunocompromised patients, outpatient sotrovimab therapy was implemented for all lung transplant recipients on February 13, 2022. Before this policy, 11 out of 16 (69%) lung transplant recipients diagnosed with COVID-19 required hospital admission, seven out of 11 required supplemental oxygen (64%) of which four (36%) required at least 5 l/min, four (36%) required at least 15 l/min or high flow nasal oxygen therapy, and one patient died. After the implementation of sotrovimab as outpatient therapy, one out of 14 patients (7%) was hospitalized (p<0.001).

### Associations with Hospitalization

We identified baseline predictors of hospitalization for these patients in an exploratory way using a univariate logistic regression analysis. This showed that a higher age, a lower IgG titer < 300 BAU/mL, being a lung transplant recipient, a higher number of comorbidities, and a higher CFS, were significantly associated with hospitalization (Table 3). The number of hospitalizations was too low to allow identification of predictors in a multivariate regression analysis.

**Table 3:** Univariate logistic regression analysis.

| Independent variable | p-value | OR | 95% CI |  |
| --- | --- | --- | --- | --- |
|  |  |  | lower | CI upper |
| Sex (male/female) | 0.43 | 0.69 | 0.27 | 1.7 |
| Age (year) | 0.00036 | 1.1 | 1.0 | 1.1 |
| Ethnicity | 0.12 | 2.1 | 0.81 | 5.6 |
| CKD (yes vs no) | 0.090 | 2.3 | 0.88 | 5.9 |
| Use of MMF (yes vs no) | 0.058 | 2.8 | 0.96 | 8.3 |
| Fully vaccinated (yes vs no) | 0.091 | 0.35 | 0.10 | 1.2 |
| Boosted (yes vs no) | 0.11 | 2.7 | 0.79 | 9.0 |
| IgG titer (BAU/mL) | 0.091 | 0.99 | 0.99 | 1.0 |
| Being an adequate responder ($\geq 300$ BAU/mL) | 0.0064 | 0.053 | 0.0060 | 0.44 |
| Being a KTR (yes vs no) | 0.88 | 1.1 | 0.42 | 2.8 |
| Being a LUTR (yes vs no) | <0.000010 | 16 | 4.7 | 53 |
| Obesity (yes vs no) | 0.22 | 2.6 | 0.57 | 12 |
| Number of comorbidities (0-5) | 0.00065 | 2.1 | 1.4 | 3.2 |
| Frailty score (1-9) | 0.00092 | 1.8 | 1.3 | 2.6 |

Dependent variable is hospitalization versus no hospitalization.
Abbreviations: OR= odds ratio , CI= confidence interval , CKD= chronic kidney disease (eGFR < 60 mL/min/1.73m<sup>2</sup>), MMF= mycophenolate mofetil,
LUTR= lung transplant recipient, KTR= kidney transplant recipients, Obesity: if BMI > 35 kg/m<sup>2</sup>

## DISCUSSION

Compared with previous SARS-CoV-2 variants, infection with Omicron results in fewer hospital admissions and a lower mortality rate [12]. However, little is known about the course of disease of Omicron in immunocompromised patients. Here, we describe the clinical characteristics, morbidity and mortality in 114 severely immunocompromised patients with COVID-19, when Omicron BA.1 was dominant in the Netherlands. Of all patients, 89% had previously received three mRNA vaccinations and 12% already received a fourth vaccination. State of the art COVID-19 therapy was available to all. In this specific context, the overall mortality of 1% is in sharp contrast to reported mortality rates ranging from 14% to 20% in hospitalized SOTRs after infection with previous VOCs [21]. However, the morbidity remained substantial as 20% required hospital admission for a median duration of 11 days. Furthermore, 25% of patients had not yet fully recovered at the end of the follow-up period of 68 days after symptom onset.

Univariate logistic regression analysis showed that several variables were associated with an increased risk of hospitalization. Not unexpected these were: a higher age, a higher number of comorbidities, and a higher CFS. A SARS-CoV-2 IgG spike antibody titer <300 BAU/mL at diagnosis (despite that 89% of all patients had previously received three mRNA vaccinations) and being a lung transplant recipient, were other relevant factors. Associations with hospital admission should be interpreted with caution, as the number of endpoints was small, the number of potential risk factors high, and therefore a comprehensive multivariate analysis was not possible. Lung transplant recipients were clearly at increased risk with a high percentage of hospital admissions (69%). This confirms observations during the circulation of previous variants and can have multiple explanations including the use of higher doses of immunosuppressive therapy, the respiratory nature of their condition, and transplanted lungs suffering more from the recipient’s immune response [22–25]. In addition, nine out of 43 (21%) kidney transplant recipients required hospital admission. The observations in lung –and kidney transplant recipients were in sharp contrast to the disease course observed in 17 heart and 18 liver transplant recipients, as none of them required hospitalization and none received sotrovimab therapy as outpatient. Of the other 14 immunocompromised patients (on anti-CD20 therapy or alloHSCT), two required hospital admission and three eventually received outpatient treatment with sotrovimab for prolonged symptoms and ongoing viral replication (with low cycle threshold PCR values).

In the pre-Omicron era, treatment with casirivimab/imdevimab of patients who were (SARS-CoV-2 IgG spike) seronegative at time of hospital admission, was associated with a 20% lower overall mortality [26]. Although a similar study was not performed with other monoclonal antibodies in seronegative patients, we considered it sufficiently likely that sotrovimab would have a similarly beneficial effect in immunocompromised patients with a low or negative SARS-CoV-2 IgG spike antibody titer. Therefore, the hospital COVID-19 treatment guideline recommended the off-label use of sotrovimab for these patients as soon as the drug became available. Strikingly, despite that 78% of the hospitalized patients had been fully vaccinated, the majority (70%) was still seronegative and 92% had a titer of <300 BAU/mL. Of the 23 hospitalized patients, 17 received sotrovimab. While 16 patients could be discharged alive without the need for mechanical ventilation, one patient with multiple comorbidities and a frailty score of seven, in whom mechanical ventilation was deliberately not initiated, died. The decision to be restrictive with sotrovimab as outpatient therapy, had to do with the uncertainty of benefit of sotrovimab for Omicron, in a setting where the vaccination coverage was 90%. However, the high hospital admission rate observed in lung transplant recipients resulted in a policy change, where sotrovimab outpatient therapy was implemented for all lung transplant recipients, immediately after COVID-19 diagnosis. Before this policy change, 11 out of 16 (69%) lung transplant recipients, diagnosed with Omicron, required hospital admission. After this policy change, 14 lung transplant recipients received sotrovimab in an outpatient setting, of which only one patient (7%) required hospital admission. This patient was hospitalized with both diverticulitis and COVID-19.

Our study has several limitations. Firstly, it is possible that the observed morbidity is overestimated since we cannot exclude that mild COVID-19 cases remained undiagnosed. However, test locations are easily accessible across the Netherlands, self-tests are available at very low costs, and the importance of testing in these vulnerable patients is constantly emphasized by their health care workers. In addition, our cohort is relatively small, with the majority being a SOTR, and therefore it represents a rather small part of the diverse immunocompromised patient population. Studies on the burden of infection with Omicron in other prevalent patient groups, including those treated for a chronic lymphocytic leukemia, multiple myeloma or other T or B-cell diseases, are therefore needed. Finally, a part of the hospital admissions could have been prevented by the use of sotrovimab in the outpatient setting. Unfortunately, with BA.2 now being the dominant variant in the Netherlands, sotrovimab is unlikely to be of benefit as its in vitro activity to this variant is limited [9,11].

In conclusion, in a highly vaccinated population of immunocompromised patients, we observed a very low mortality from COVID-19 caused by Omicron. The morbidity however, continues to be very substantial despite Omicron BA.1 being the dominant variant. In particular lung– and kidney transplant recipients are likely to benefit from early treatment for COVID-19. However, the recently reported reduced or complete absence of activity of sotrovimab against the BA.2 variant, will require the use of other monoclonal antibodies or directly acting antiviral drugs like nirmatrelvir/ritonavir, once they become broadly available.

## Data Availability

All data produced in the present study are available upon reasonable request to the authors

