## Supplemental text for "Clinical characteristics and outcome of immunocompromised patients with COVID-19 caused by the Omicron variant: a prospective observational study"

### Online supplement

**Single-nucleotide polymorphisms (SNPs) analysis**

The combination of SNP PCRs is able to discriminate the following specific mutations in spike: K417K/N, S371S/L/F and S373S/P. The results from the SNP PCRs were interpreted in the context of the molecular epidemiology of SARS-CoV-2 variants during the study period in the Netherlands. Results were interpreted as follows: 1. K417N + S371L + S373P = highly indicative of Omicron variant, BA.1 sublineage. 2. K417N + S371F + S373P = highly indicative of Omicron variant, BA.2 sublineage. 3. K417N + any other combination = highly indicative of Omicron variant, sublineage unknown. 4. K417K + S371S + S373S = highly indicative of Delta variant, sublineage unknown.

**Standard Immunosuppressive Regime**

All solid organ transplant recipients have received induction therapy with either basiliximab 20 mg intravenously on day 0 and 4 after transplantation (kidney, lung and liver), or methylprednisolone on day 0 (liver), or anti-thymocyte globulin (heart) on day 0 and 1 after transplantation. Standard maintenance immunosuppression included corticosteroids (dose according to weight and time after transplant, 0.05-0.1 mg/kg/day) or fixed dose in heart and liver transplant recipients, tacrolimus with trough levels in between 4-8 µg/L (liver and kidney), 7-10 µg/L (heart and lung), and mycophenolate mofetil. Steroids were tapered at four, six and 12 months respectively (kidney, lung, heart and liver), if feasible. For reasons of toxicity, graft rejection or tolerance, alternative immunosuppressive schemes could be administered, such as mTOR inhibitors or azathioprine. The "non-solid organ transplant patient group" consisted of patients treated with rituximab within the last six months for an underlying hematological or autoimmune disease as well as patients after allogeneic hematopoietic stem cell transplantation on immunosuppressive therapy for prevention or treatment of graft versus host disease.
